# Real-World Evaluation of an Automated Algorithm to Detect Patients with Potentially Undiagnosed Hypertension in an Ethnically Diverse, Large Health System in Hawaiʻi

**DOI:** 10.1101/2023.06.16.23291529

**Authors:** Mika D. Thompson, Yan Yan Wu, Blythe Nett, Lance K. Ching, Hermina Taylor, Tiffany Lemmen, Tetine L. Sentell, Meghan D. McGurk, Catherine M. Pirkle

## Abstract

**Objective:** This real-world evaluation considers an algorithm designed to detect patients with potentially undiagnosed hypertension, receiving routine care, in a large health system in Hawaiʻi. It quantifies patients identified as potentially undiagnosed with hypertension, summarizes the individual, clinical, and health system factors associated with undiagnosed hypertension, and examines if the COVID-19 pandemic impacted detection.

**Methods:** We analyzed the electronic health records (EHR) of patients treated across 6 clinics from 2018-2021. We calculated total patients with potentially undiagnosed hypertension and compared patients flagged for undiagnosed hypertension to those with diagnosed hypertension and to the full patient panel across individual characteristics, clinical and health system factors (e.g., clinic of care), and timing. Modified Poisson regression was used to calculate crude and adjusted risk ratios.

**Results:** Among the eligible patients (N=13,364), 52.6% had been diagnosed with hypertension, 2.7% were flagged as potentially undiagnosed, and 44.6% had no evidence of hypertension. Factors associated with a higher risk of potentially undiagnosed hypertension included: individual characteristics (ages 40-84 compared to 18-39 years), clinical (lack of diabetes diagnosis) and health system factors (clinic site and being a Medicaid versus a Medicare beneficiary), and timing (readings obtained after the COVID-19 Stay-At-Home Order in Hawaiʻi).

**Conclusions:** This evaluation provided evidence that a clinical algorithm implemented within a large health systems’s EHR could detect patients in need of follow-up to determine hypertension status, and it identified key individual characteristics, clinical and health system factors, and timing considerations that may contribute to undiagnosed hypertension among patients receiving routine care.

## INTRODUCTION

Hypertension affects half of the adult population in the United States (US),^1^ with the number of adults with hypertension worldwide doubling over the last three decades.^2^ In the US, hypertension presents a substantial financial burden, with the total associated medical costs estimated at $131 billion annually.^3^ Hypertension is an influential component to many life-threatening cardiovascular outcomes, such as stroke and heart failure,^4^ and is strongly associated with morbidity and premature mortality.^5^ In 2019, the Center for Disease Control (CDC) listed hypertension as a primary or contributing cause of over half a million deaths in the US.^6^ Despite robust clinical standards for diagnosing and managing hypertension,^7^ undiagnosed hypertension remains a significant public health issue around the world.^8^ In the US, an estimated 11 million people (5.9%) who have a usual source of care have undiagnosed hypertension.^9^ Those without a usual source of care are even more likely to have an undiagnosed chronic disease.^10^

Growing evidence demonstrates that even patients with access to regular health care can go undiagnosed.^11, 12^ For example, in a nationally representative analysis of undiagnosed hypertension in the US, over 80% had health insurance and/or a usual source of care, and were termed as “hiding in plain sight.”^12^ System-wide interventions to identify undiagnosed cases of hypertension within health care settings could improve clinical care and reduce the economic and health consequences of undiagnosed and unmanaged hypertension.

Automated algorithms, a health information technology (HIT), may provide an opportunity to identify patients with undiagnosed hypertension to initiate management, improve health outcomes, and reduce costs.^13^ These algorithms can identify potential cases of a condition based on abnormal test results found in patients’ electronic health records (EHR) and flag patients that fit a clinical definition, but lack a record of diagnosis or management of the condition. EHR algorithms have been used to identify patients with many potentially undiagnosed chronic conditions, such as diabetes^14–16^ and hypertension.^8, 9, 12, 17, 18^

The potential of using EHRs in ambulatory care to identify undiagnosed hypertension is also well-demonstrated.^12, 18, 19^ Hypertension is a condition particularly amenable to the application of automated EHR algorithms given the ubiquity of blood pressure assessments during routine clinical encounters. Despite the use of automated EHR algorithms in surveillance research and the promising potential of such interventions in clinical settings to improve the detection and management of hypertension,^9^ limited empirical research tests the application of such technology in real-world clinical settings.^20–22^

Hawaiʻi-based health systems offer a unique opportunity to assess the use of automated algorithms within a population largely comprised of understudied and underrepresented groups, including Native Hawaiians (NH), Other Pacific Islanders (OPI), and diverse Asian groups.^23^ Moreover, NH and OPI often lack representation in medical school and among the physician population in Hawaiʻi, further emphasizing the need to address health disparities and ensure equitable healthcare services for these communities.^24, 25^ Hawaiʻi adults have the highest predicted prevalence of undiagnosed hypertension in the US (6.5%), with several predominant racial and ethnic groups experiencing a disproportionate burden of diagnosed hypertension.^26, 27^ Implicit biases on the part of providers may contribute to unequal care and treatment of minority groups.^28–30^ While there are evident disparities in diagnosed hypertension in Hawaiʻi, it remains unclear whether similar disparities exist for undiagnosed hypertension. In other words, are certain groups more likely to be missed than others? Hawaiʻi provides an ideal setting to examine this question, as over 90% of the population has insurance coverage, greatly reducing a major contributor to undiagnosed hypertension.^31^

We conducted a real-world evaluation^32^ of an automated algorithm designed to identify patients with undiagnosed hypertension implemented within the Queen’s Clinically Integrated Physician Network (QCIPN), a physician organization within the Queen’s Health System (QHS). The objectives were to: (1) determine the number of patients flagged as undiagnosed hypertension by the algorithm, (2) summarize associated individual characteristics, clinical, and health system factors, and (3) assess whether the COVID-19 pandemic affected detection. The latter objective emerged as some of the analytic period overlapped with the early stages of the pandemic.

## METHODS

### Design and Setting

This quality improvement evaluation examined the de-identified EHR data of patients from 6 QCIPN clinics on Oʻahu. QHS serves a patient population that is approximately 25% NH and OPI, 20% White, 20% Japanese, and 17% Filipino. Automated algorithms were developed by the Hawaiʻi Department of Health (HDOH). The algorithm was supported using pilot funding from two CDC cooperative agreements (1305 and 1815). An evaluation was implemented to assess the algorithm function in practice. QHS administrators selected 6 clinics to apply the automated algorithm at scale. These clinics provide primary care and some specialty services. De-identified patient data from July 2, 2018 to July 1, 2021 was provided. Use of these de-identified secondary data for evaluation was deemed non-human subjects research by the HDOH and University of Hawai‘i Institutional Review Boards per the Revised Common Rule of 2018.

### Intervention

The algorithm was developed to be evidence-based and grounded in clinical recommendations from the CDC and Centers for Medicare and Medicaid Services (CMS) Million Hearts Initiative (Supplementary Figure S1). In brief, the algorithm pulls patient’s EHR data in a population health management system, searches for hypertension diagnoses, and screens for in-office blood pressure measurements. Through these steps, different prompts are presented to the patient’s primary care provider with recommended actions for care. QHS implemented the algorithms. The algorithms were applied at the population health management level to resolve interoperability issues that occur when providers or services within a health system use different EHR software. Patients flagged by the algorithm for potentially undiagnosed hypertension were listed in a registry created by a software called Population Builder. External analysis of patient data, including those flagged by the algorithm, was conducted by researchers at the University of Hawaiʻi.

### Population

Eligible patients were 18 years or older and had at least one encounter with a physician at 1 of the 6 clinics during the three-year period. This was done to ensure that the sample population consisted of patients actively engaged in healthcare, rather than those lost-to-follow-up or inactive. The exclusion criteria were: (1) those with resolved hypertension (i.e., hypertension that is no longer on the problem list, and lacked two systolic and diastolic blood pressure measurements ≥140/90 in the past three years or one measurement ≥160/100 in the past year), (2) those who screened for hypertension within the one year prior to the end of the measurement period, but a diagnosis was deemed inappropriate, (3) those with an active diagnoses of end-stage renal disease (ascertained through EHR using ICD-9 codes: 585.5, 585.6, V42.0, V45.1, and V56, and ICD-10 codes: N18.5, N18.6, Z49, Z94.0, Z91.15, and Z99.2), (4) those with no hypertension diagnosis but were taking hypertension medication, and (5) those that were deceased by the end of the measurement period.

### Outcomes

Three groups were derived from patients’ EHR:

1. Physician-diagnosed hypertension: These cases were unresolved cases of hypertension derived from the patients’ active problem list identified through ICD-9 codes 401- 405 and 410.9, and ICD-10 codes I10-I15 and I21.3.
2. Potentially undiagnosed hypertension: This was the primary outcome measure. Cases were defined as having: two continuously measured systolic or diastolic blood pressure measurements ≥140/90 in the past three years, or one measurement ≥160/100 in the past year and no hypertension diagnosis on the patient’s active problem list.
3. No indication of high blood pressure: These cases consisted of patients that were not flagged by the algorithm based on the above criteria and did not have hypertension on their active problem list.

### Covariates

We compared potentially undiagnosed cases to diagnosed ones across individual characteristics, clinical and health system factors, and timing. The covariates described below were selected based on evidence in the literature that these factors may be associated with hypertension diagnosis. Measures were extracted from patients’ EHRs.

#### Individual Characteristics

These included patient demographics: age in years (at extraction date) grouped into categories (18-39; 39-65; 65-85; or 85-106), sex (male/female), race and ethnicity (East Asian, Filipino, NH, Other, OPI, or White). Race and ethnicity was self-reported by patients and recorded in their EHR. Coding for the multiple response race and ethnicity variable was derived using the following rules: (1) if NH was one of the multiple ethnicities listed, “Native Hawaiian” was coded, (2) if a non-White ethnicity was listed with a White ethnicity, the non-White ethnicity was coded, and (3) if there was more than one non-White ethnicity listed, “East Asian” was coded because all patients in the sample endorsing more than one non-White ethnicity (N=4) were multiple East Asian ethnicities (e.g., Japanese and Korean). Patients classified as “Other” race and ethnicity included American Indian, Alaska Native, Black, and Hispanic. A potential limitation of using this method to create mutually exclusive race and ethnicity categories is the creation of groups with substantial proportions of multiracial patients, especially given Hawaiʻi’s large multiracial population.^23^

Patient behavioral measures included any tobacco use (yes/no; ICD-10 codes F17, F17.20-F17.22, F17.29, Z72.0, Z57.31, Z72.0, Z77.22, and Z87.891) and any alcohol use (yes/no) extracted from social history.

#### Clinical and Health System Factors

These measures reflected health concerns documented in the EHR, as well as clinic site and insurer. Measures included the most recent categorical body mass index (underweight, <18.5; normal, (18.5, 25.0); overweight, (25.0, 30.0); or obese, ≥30.0) and any mental health conditions (yes/no; ICD-10 codes F20-F29, F30-F39, F40-F48, and F60-F69). A five-category indicator for prediabetes and diabetes was also included. The first two categories reflected a diagnosis of prediabetes or diabetes in the EHR (ICD-10 codes E08, E09, E10, E11, E13, and R73.03). We used the most recently available A1C value to categorize the remaining patients. Those with an A1C below 5.7 were categorized as without diabetes. Those with an A1C of 5.7 or greater, but without a diagnosis, were considered potential cases of prediabetes or diabetes.^33^ Finally, there was a group of patients that lacked any diabetes diagnosis or A1C value in three years.

Insurer was categorized into Medicare, Blue Cross Blue Shield (BCBS), Medicaid, or Other (e.g., uninsured, Department of Defense). Clinic site was anonymized and each was assigned a number.

#### Timing

To assess the possible effect of the pandemic on risk of potentially undiagnosed hypertension, patients’ records were split into two time periods: before and after Hawaiʻi’s first statewide Stay-At-Home Order, henceforth referred to as “the Order”.^34^ This emergency proclamation, requiring all Hawai‘i residents to remain home for all non-essential activities, took effect on March 25, 2020. Just before this declaration, CMS announced that non-essential medical procedures should be delayed to preserve medical supplies.^35^ Decisions on which services were non-essential were left to state and local officials. Immediately following the Order, healthcare services were severely restricted, with many non-COVID-19-related appointments postponed or cancelled, including routine and preventative care.^36, 37^ The clinics in this evaluation reduced in-person appointments during the post-pandemic period. The timing variable roughly captured whether the automated algorithm flagged patients for potentially undiagnosed hypertension before or after the Order.

### Statistical Methods

Sample characteristics were summarized by frequency and percentage across patient groups according to the total sample, those with diagnosed hypertension, and those flagged with potentially undiagnosed hypertension.

Utilizing modified Poisson regression with a robust error variance,^38^ we calculated the crude and adjusted relative risk (RR) and 95% confidence intervals (95%CI) for patients flagged with potentially undiagnosed hypertension by sample characteristics. For the adjusted RR estimates, we created three multivariate models. The first examined only the individual characteristics. The second added the clinical and health system factors, while the third added the COVID-19 timing variable. Adjustment sets for the multivariate models reflected the modelling strategy. Analyses were conducted using R version 4.1.2 with integrated development environment RStudio version 1.4.1106.

## RESULTS

### Analytic Sample

Figure 1 presents the derivation of the analytic sample. There were 14,497 eligible patients, 7,032 had a hypertension diagnosis, and 7,465 did not. Among the 7,465 with no diagnosis, 788 were excluded based on exclusion criteria and 345 were excluded due to missing covariates, resulting in a final analytic sample of 13,364 patients. Of these, the automated algorithm flagged 366 (2.7%) for potential undiagnosed hypertension. An additional 7032 (52.6%) had physician-diagnosed hypertension and 5,966 (44.6%) had no indication of hypertension.

**Figure 1.**
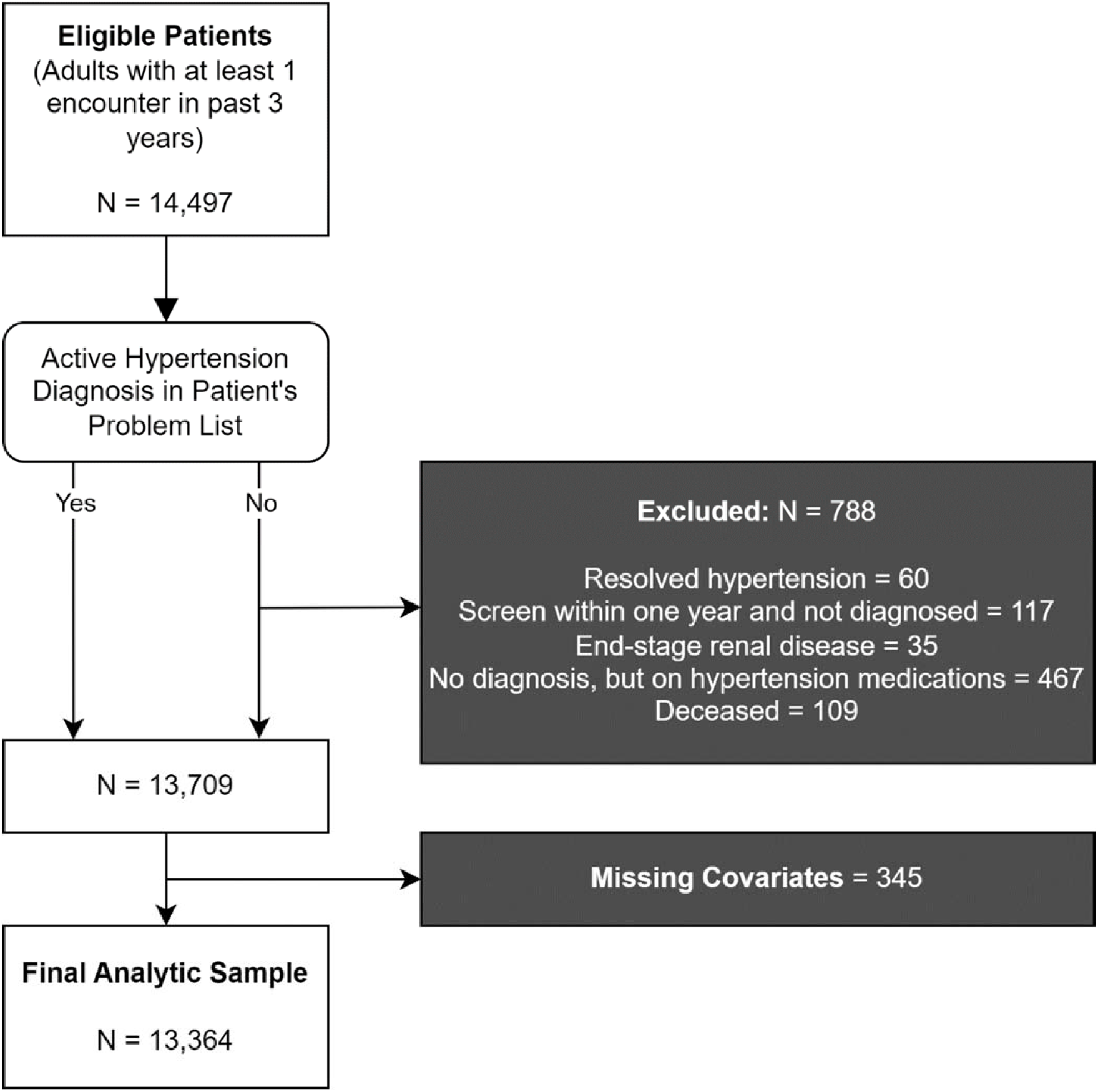
Sample Derivation using the QCIPN Algorithm

### Patient Characteristics by Hypertension Diagnosis Status

Table 1 shows the distribution of individual, clinical, and health system factors, whether the patient was flagged for potential undiagnosed hypertension, and the unadjusted associations for those flagged according to the covariates examined.

**Table 1.**
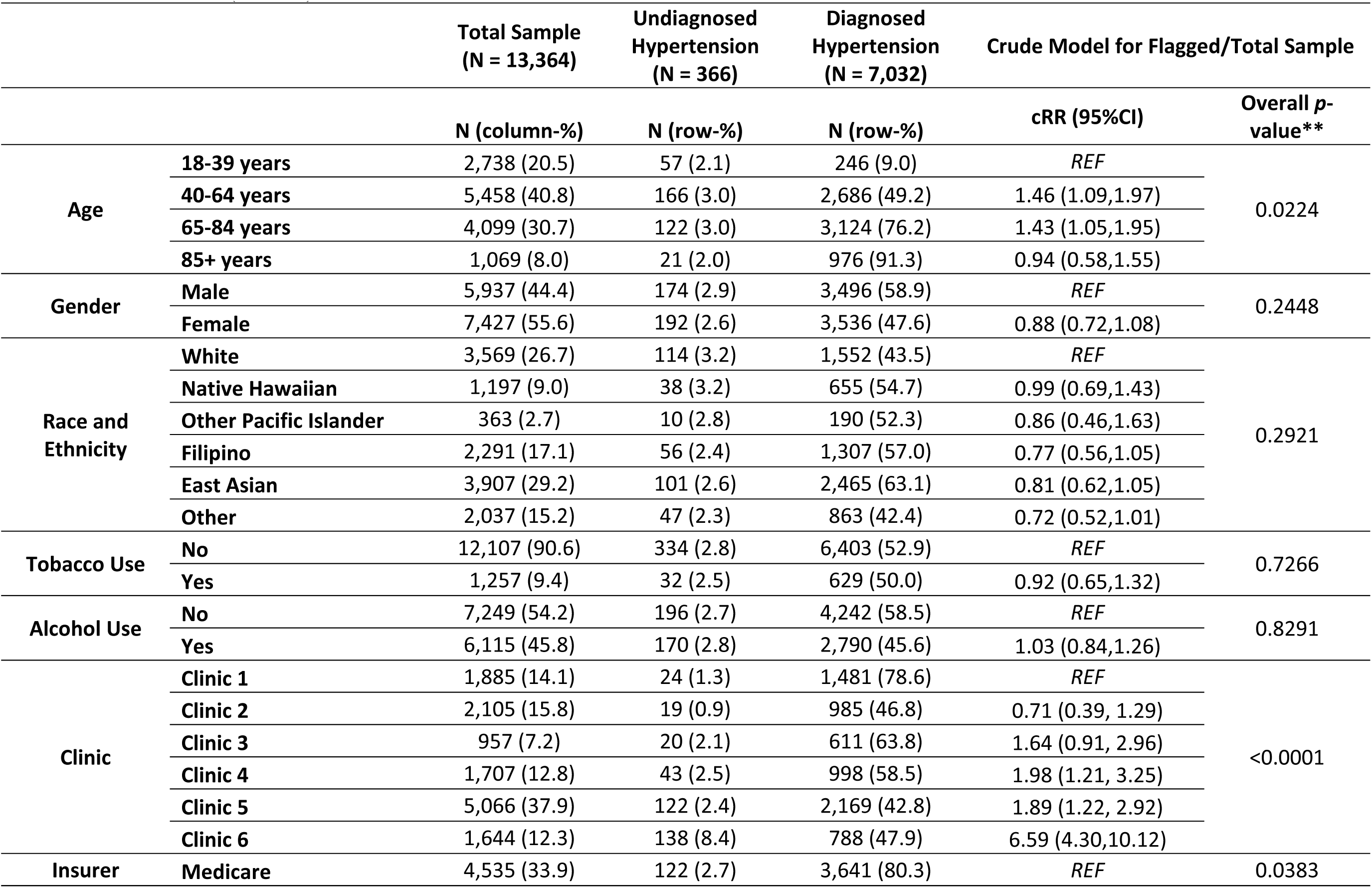
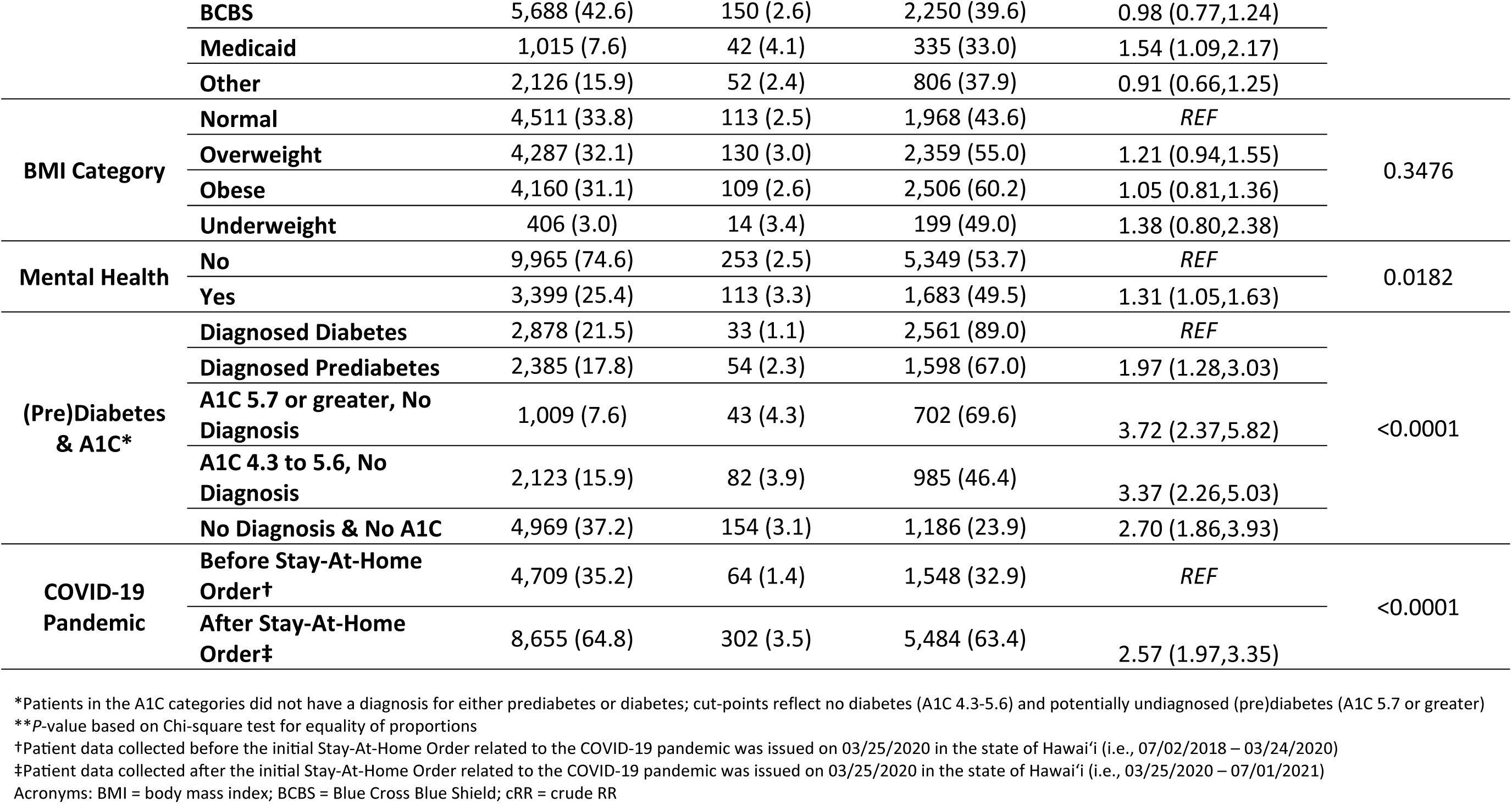
Sample Characteristics by Undiagnosed Hypertension and Diagnosed Hypertension, with Crude Relative Risk (RR) and 95% Confidence Intervals (95%CI)

|  |  | Total Sample<br>(N = 13,364) | Undiagnosed<br>Hypertension<br>(N = 366) | Diagnosed<br>Hypertension<br>(N = 7,032) | Crude Model for Flagged/Total Sample |  |
| --- | --- | --- | --- | --- | --- | --- |
|  |  | N (column-%) | N (row-%) | N (row-%) | cRR (95%CI) | Overall p-value** |
| Age | 18-39 years | 2,738 (20.5) | 57 (2.1) | 246 (9.0) | REF | 0.0224 |
|  | 40-64 years | 5,458 (40.8) | 166 (3.0) | 2,686 (49.2) | 1.46 (1.09,1.97) |  |
|  | 65-84 years | 4,099 (30.7) | 122 (3.0) | 3,124 (76.2) | 1.43 (1.05,1.95) |  |
|  | 85+ years | 1,069 (8.0) | 21 (2.0) | 976 (91.3) | 0.94 (0.58,1.55) |  |
| Gender | Male | 5,937 (44.4) | 174 (2.9) | 3,496 (58.9) | REF | 0.2448 |
|  | Female | 7,427 (55.6) | 192 (2.6) | 3,536 (47.6) | 0.88 (0.72,1.08) |  |
| Race and Ethnicity | White | 3,569 (26.7) | 114 (3.2) | 1,552 (43.5) | REF | 0.2921 |
|  | Native Hawaiian | 1,197 (9.0) | 38 (3.2) | 655 (54.7) | 0.99 (0.69,1.43) |  |
|  | Other Pacific Islander | 363 (2.7) | 10 (2.8) | 190 (52.3) | 0.86 (0.46,1.63) |  |
|  | Filipino | 2,291 (17.1) | 56 (2.4) | 1,307 (57.0) | 0.77 (0.56,1.05) |  |
|  | East Asian | 3,907 (29.2) | 101 (2.6) | 2,465 (63.1) | 0.81 (0.62,1.05) |  |
|  | Other | 2,037 (15.2) | 47 (2.3) | 863 (42.4) | 0.72 (0.52,1.01) |  |
| Tobacco Use | No | 12,107 (90.6) | 334 (2.8) | 6,403 (52.9) | REF | 0.7266 |
|  | Yes | 1,257 (9.4) | 32 (2.5) | 629 (50.0) | 0.92 (0.65,1.32) |  |
| Alcohol Use | No | 7,249 (54.2) | 196 (2.7) | 4,242 (58.5) | REF | 0.8291 |
|  | Yes | 6,115 (45.8) | 170 (2.8) | 2,790 (45.6) | 1.03 (0.84,1.26) |  |
| Clinic | Clinic 1 | 1,885 (14.1) | 24 (1.3) | 1,481 (78.6) | REF | <0.0001 |
|  | Clinic 2 | 2,105 (15.8) | 19 (0.9) | 985 (46.8) | 0.71 (0.39, 1.29) |  |
|  | Clinic 3 | 957 (7.2) | 20 (2.1) | 611 (63.8) | 1.64 (0.91, 2.96) |  |
|  | Clinic 4 | 1,707 (12.8) | 43 (2.5) | 998 (58.5) | 1.98 (1.21, 3.25) |  |
|  | Clinic 5 | 5,066 (37.9) | 122 (2.4) | 2,169 (42.8) | 1.89 (1.22, 2.92) |  |
|  | Clinic 6 | 1,644 (12.3) | 138 (8.4) | 788 (47.9) | 6.59 (4.30,10.12) |  |
| Insurer | Medicare | 4,535 (33.9) | 122 (2.7) | 3,641 (80.3) | REF | 0.0383 |
|  | **BCBS** | 5,688 (42.6) | 150 (2.6) | 2,250 (39.6) | 0.98 (0.77,1.24) |  |
|  | **Medicaid** | 1,015 (7.6) | 42 (4.1) | 335 (33.0) | 1.54 (1.09,2.17) |  |
|  | **Other** | 2,126 (15.9) | 52 (2.4) | 806 (37.9) | 0.91 (0.66,1.25) |  |
| **BMI Category** | **Normal** | 4,511 (33.8) | 113 (2.5) | 1,968 (43.6) | REF | 0.3476 |
|  | **Overweight** | 4,287 (32.1) | 130 (3.0) | 2,359 (55.0) | 1.21 (0.94,1.55) |  |
|  | **Obese** | 4,160 (31.1) | 109 (2.6) | 2,506 (60.2) | 1.05 (0.81,1.36) |  |
|  | **Underweight** | 406 (3.0) | 14 (3.4) | 199 (49.0) | 1.38 (0.80,2.38) |  |
| **Mental Health** | **No** | 9,965 (74.6) | 253 (2.5) | 5,349 (53.7) | REF | 0.0182 |
|  | **Yes** | 3,399 (25.4) | 113 (3.3) | 1,683 (49.5) | 1.31 (1.05,1.63) |  |
| **(Pre)Diabetes & A1C\*** | **Diagnosed Diabetes** | 2,878 (21.5) | 33 (1.1) | 2,561 (89.0) | REF | <0.0001 |
|  | **Diagnosed Prediabetes** | 2,385 (17.8) | 54 (2.3) | 1,598 (67.0) | 1.97 (1.28,3.03) |  |
|  | **A1C 5.7 or greater, No Diagnosis** | 1,009 (7.6) | 43 (4.3) | 702 (69.6) | 3.72 (2.37,5.82) |  |
|  | **A1C 4.3 to 5.6, No Diagnosis** | 2,123 (15.9) | 82 (3.9) | 985 (46.4) | 3.37 (2.26,5.03) |  |
|  | **No Diagnosis & No A1C** | 4,969 (37.2) | 154 (3.1) | 1,186 (23.9) | 2.70 (1.86,3.93) |  |
| **COVID-19 Pandemic** | **Before Stay-At-Home Order†** | 4,709 (35.2) | 64 (1.4) | 1,548 (32.9) | REF | <0.0001 |
|  | **After Stay-At-Home Order‡** | 8,655 (64.8) | 302 (3.5) | 5,484 (63.4) | 2.57 (1.97,3.35) |  |

\*Patients in the A1C categories did not have a diagnosis for either prediabetes or diabetes; cut-points reflect no diabetes (A1C 4.3-5.6) and potentially undiagnosed (pre)diabetes (A1C 5.7 or greater)
\*\*P-value based on Chi-square test for equality of proportions
†Patient data collected before the initial Stay-At-Home Order related to the COVID-19 pandemic was issued on 03/25/2020 in the state of Hawai'i (i.e., 07/02/2018 – 03/24/2020)
‡Patient data collected after the initial Stay-At-Home Order related to the COVID-19 pandemic was issued on 03/25/2020 in the state of Hawai'i (i.e., 03/25/2020 – 07/01/2021)
Acronyms: BMI = body mass index; BCBS = Blue Cross Blue Shield; cRR = crude RR

Most patients in the analytic sample were 40 years of age or older (79.5%). The analytic sample skewed slightly female. East Asian was the most prevalent racial and ethnic group (29.2%), followed by White (26.7%) and Filipino (17.1%). Tobacco use was 9.4%, while about half of patients reported alcohol use. About a quarter of patients were diagnosed with diabetes and 17.8% diagnosed with prediabetes. A large proportion of patients were sampled from Clinic 5 (37.9%), followed by Clinic 2 (15.8%) and Clinic 1 (14.1%). Most patients were insured through Medicare or BCBS, with a small percentage using Medicaid (7.6%). Finally, the majority of patients’ readings were recorded after the Order (64.8%).

### Potentially Undiagnosed versus Diagnosed Hypertension

The proportion of patients with potentially undiagnosed hypertension significantly differed by age group (*p*=0.0224), clinic (*p*<0.0001), insurer (*p*=0.0383), mental health (*p*=0.0182), (pre)diabetes & A1C (*p*<0.0001), and whether readings were obtained before or after the Order (*p*<0.0001). Those with diagnosed prediabetes were more likely to have potentially undiagnosed hypertension relative to those with diagnosed diabetes. However, the percentage of those in this group (2.3%) was less than that of potentially undiagnosed hypertension patients in the total sample (2.7%). Those with evidence of prediabetes (A1C 4.3- 5.6, No Diagnosis) or diabetes (A1C ≥ 5.7, No Diagnosis), based on A1C value, were also more likely to be flagged for undiagnosed hypertension. Those with no A1C records and no diagnosis of (pre)diabetes (No Diagnosis & No A1C) were also more likely to be flagged compared to patients with diabetes; however, the percent of potentially undiagnosed hypertension among those with no A1C record and no (pre)diabetes diagnosis (3.1%) was similar to the percentage in the full sample. Over 8% of all patients at Clinic 6 were flagged. Those on Medicaid (4.1%) were more likely to be flagged compared to those on Medicare (2.7%). Before the Order, only 1.4% of patients followed at these six clinics were flagged, whereas after, 3.5% of patients were flagged.

### Multivariable Risk of Potentially Undiagnosed Hypertension

Table 2 presents the adjusted relative risks for undiagnosed hypertension across individual characteristics, clinical and health system factors, and timing. When examining individual characteristics only, middle-aged (40-64) and older adults (65-84) were more likely to be flagged compared to those 18-39 years. No other individual characteristic was significantly associated with undiagnosed hypertension in the first model.

**Table 2.**
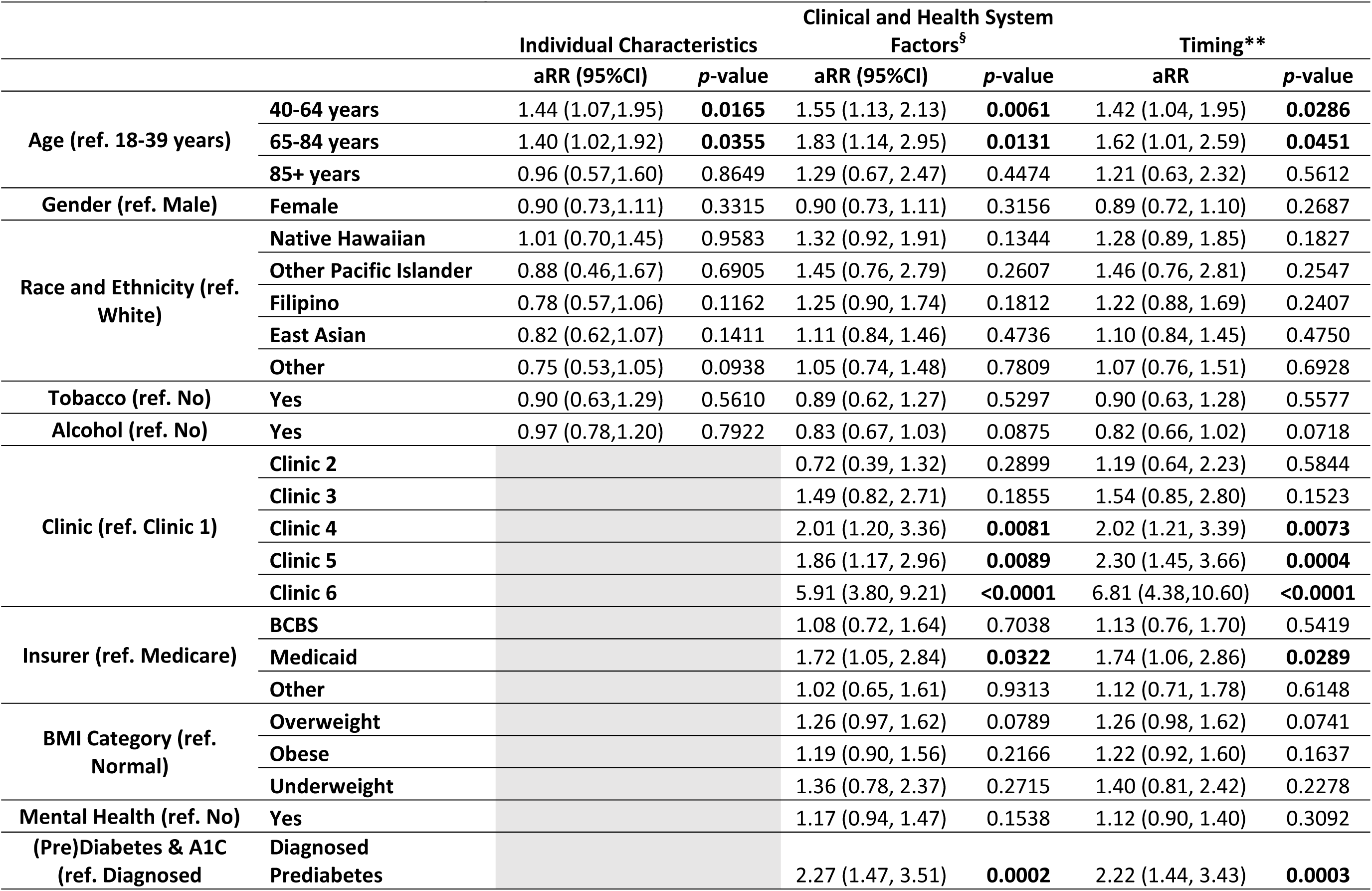
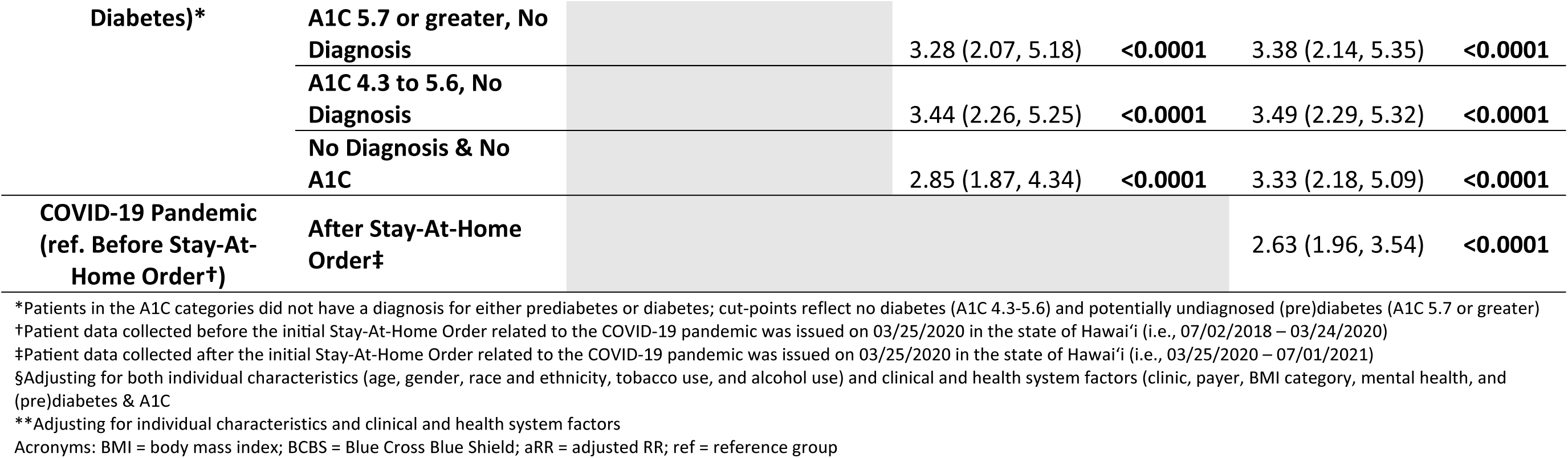
Adjusted Relative Risk (RR) and 95% Confidence Intervals (95%CI) for Undiagnosed Hypertension Across Individual Factors, Care Utilization Indicators, and Timing

The second model added clinical and health system factors. Age remained significantly associated with undiagnosed hypertension. Medicaid was significantly associated with being flagged compared to Medicare. Similar to the unadjusted models in Table 1, those with diagnosed diabetes were significantly less likely to be flagged compared to all other (pre)diabetes categories. The strongest association, as observed in the unadjusted model, was with clinic site. Those followed at Clinic 6 were nearly six times more likely to be flagged than those at Clinic 1.

The final model added the timing of the most recent reading either before or after the Order. The association with age and Medicaid remained and the association with clinic strengthened. Flagging for undiagnosed hypertension increased at all clinics after the Order, with a notable increase for Clinic 6 (3.7% to 10.7%, Supplementary Table S1). Those with diabetes remained far less likely to be flagged than any other category. The effect estimates for most of the (pre)diabetes and A1C categories changed very little between models. Finally, the risk of being flagged was significantly higher after the Order compared to before it.

## DISCUSSION

This real-world evaluation described the frequency of, and factors associated with, potentially undiagnosed hypertension as identified by an automated algorithm implemented at six clinics in a large healthcare system in Hawaiʻi. Of the 13,364 patients, we found that 2.7% had potentially undiagnosed hypertension based on blood pressure measurements in their EHR. The risk of being flagged for potentially undiagnosed hypertension significantly differed by age group, clinic, insurer, (pre)diabetes diagnosis and A1C, and whether blood pressure was measured before or after the Order.

Hypertension was prevalent among patients followed at the clinics in this sample, which principally provided primary care services. Approximately half of patients had a hypertension diagnosis. Comparatively, 30% percent of the population in Hawaiʻi self-reported hypertension in 2019.^39^ Self-report underestimates population prevalence given that hypertension awareness is consistently and substantially lower than actual prevalence.^26^ The risk of hypertension is also well known to increase with advancing age^1, 40^ and approximately 40% of patients attending these clinics were 65 or older. It is estimated that 77% percent of older adults in the US have hypertension.^41^

Just under 3% of patients followed at the six clinics were potentially undiagnosed for hypertension. This was less than the predicted prevalence of 6.5% undiagnosed hypertension in the general population of Hawaiʻi,^26^ which was expected given that our sample consisted of patients with a usual source of care and observed in the last 3 years. A study of approximately 9 million US patients comparing the number of observed hypertension cases and the number of predicted cases found that the nearly 13% of patients with hypertension may not be diagnosed.^9^ Similarly, Huguet and colleagues^31^ examined a large sample of patients from a network of community health centers across 14 US states and found that 37.3% of patients had undiagnosed hypertension, with 24.9% of cases remaining undiagnosed for the five-year study. Despite methodological differences, these estimates far exceed the percentage of potentially undiagnosed hypertension observed among patients in this sample. This difference may reflect a strong health system and/or the high health insurance coverage in Hawaiʻi.

The only individual factor associated with undiagnosed hypertension was age, but not gender or race and ethnicity. There is extensive literature showing that women are less likely to receive preventative treatment or guidance for cardiovascular disease,^42^ possibly reflecting biases in scientific literature and medical training about cardiovascular disease in women.^43^ Despite notable differences in both the risk and awareness of hypertension by race and ethnicity in the existing literature,^44, 45^ the observed differences were marginal. The disparities by race and ethnicity are often attributed to structural and social factors and cultural norms, such as healthcare access, providers’ racial and ethnic implicit bias in diagnosis and treatment, and beliefs towards seeking medical help for preventive care. ^46–48^ NH have experienced multigenerational injustice, discrimination, and cultural suppression resulting from the American overthrow and colonization of the Hawaiian Islands; these events have been linked to the substantial disparities in many health risk factors and outcomes observed today within the state.^49, 50^ However, the use of patients with recent encounters with health services may have hindered our ability to detect differences by race and ethnicity that we may have otherwise observed in the general population. Moreover, while literature on health services utilization in Hawaiʻi does indicate underutilization by NH and OPI, the very high insurance coverage in the state helps to reduce disparities associated with access to care due to lack of insurance.^51^

Several clinical and health system factors were associated with undiagnosed hypertension, most notably the service clinic. Patients at Clinic 6 were nearly seven times as likely to be flagged as those at Clinic 1. The high percentage of patients with potentially undiagnosed hypertension at Clinic 6 may be related to physician turnover. Clinic 6 experienced a significant turnover in providers during the data period; specifically, two of the four full-time providers left the clinic. Prior studies demonstrate associations between physician turnover and reductions in timely care,^52^ patient satisfaction, and organizational stability.^53^ The loss of half of the full-time providers may have resulted in delays in new provider workflow orientation and/or a lack of access to care while patients are being transitioned to new providers. Additionally, the high percentage of diagnosed hypertension and low percentage of potentially undiagnosed hypertension at Clinic 1 could be attributed to Clinic 1’s strong provider champions of panel management. Panel management, a population-based care approach where clinics proactively monitor the health of and care for all patients in their panel, not just those who come in for urgent care needs,^54^ has been associated with increased screening for and improved care for chronic conditions.^55, 56^

Medicaid beneficiaries had a higher likelihood of being flagged compared to Medicare beneficiaries. Given our statistical models, and that our sample was limited to patients with regular contact with a health provider, this finding is independent of the considered risk factors of the demographic and behavioral risk factors for hypertension, including age and race and ethnicity, as well as access to health services. This suggests that broader social factors related to Medicaid eligibility (i.e., low-income) may drive the observed disparity between Medicaid and Medicare beneficiaries regarding potentially undiagnosed hypertension, such as higher order health needs or a lack of access to healthy lifestyles.

There was also an association between different indicators of diabetes and potentially undiagnosed hypertension; most notably, those with evidence of prediabetes with no diagnosis were significantly more likely to also have potentially undiagnosed hypertension. One hypothesis is that patients with missing information on one condition may be more likely to be missing an appropriate hypertension diagnosis as well.

Lastly, significantly more individuals were flagged for undiagnosed hypertension after the first statewide Stay-At-Home Order for COVID-19. This suggests that the pandemic may have had an impact on the detection and diagnosis of hypertension. Moreover, the pandemic may have had an effect on patients’ blood pressure due to stress, fear, job loss, and other related factors. The conditions during this period, such as Hawaiʻi’s disproportionately high unemployment rate,^57^ increases in mental health issues, including depression, suicidality, isolation, likely contributing to the high blood pressure rates.^58–60^ Furthermore, during the pandemic routine medical appointments and procedures were restricted,^35, 61^ and people delayed care out of fears of contracting COVID-19.^62^ As a result, it is possible that those who were seeking medical attention had more urgent health needs, which took priority over diagnosis of hypertension.

This evaluation of a quality improvement intervention has several strengths. First, even though data came from only six clinics, we had a large enough sample from which to examine associations with potentially undiagnosed hypertension, which was a relatively rare event. Furthermore, the sample was ethnically diverse, which may provide insights that can be generalized to other health systems serving diverse populations. The algorithm assessed by this evaluation was carefully crafted and received strong buy-in from QHS, which demonstrates the potential for wider implementation. Conducting this evaluation in Hawaiʻi, a state with high health insurance coverage, greatly reduced the potential for selection bias based on financial access to care. Lastly, this work assesses the performance of an EHR algorithm in a real-world clinical setting.

Our evaluation has limitations. The data did not include how often patients used healthcare services, which could affect detection of undiagnosed hypertension. It is unclear whether the number of visits varied differentially by the considered factors. All variables were derived from an EHR; as a result, we could not follow up with flagged patients to confirm undiagnosed hypertension. Selection bias through health care access/utilization may affect the generalizability of our findings. As mentioned, Hawaiʻi’s large multiracial population (nearly 25%)^23^ may have resulted in substantial proportions of multiracial patients and limited examination of differences by race and ethnicity. Moreover, the collection of information on race and ethnicity is complex and challenging for several inherent reasons, including a lack of standard definitions, differences between the constructs of race and ethnicity, and the influence of acculturation and assimilation, among others.^63^ Lastly, we could not account for masked hypertension, a hypertension subtype where a patient has normal blood pressure readings in-office while having hypertensive readings out-of-office.^64, 65^ The prevalence of masked hypertension ranges between 10-17% using daytime measurements; however, the prevalence may be far greater when accounting for daytime, nighttime, and ambulatory measurements.^66^

It is important to acknowledge that the findings from this evaluation pertain to active patients with at least one physician encounter during the three-year study. Our sample, thus, consisted only of patients with access to, and recent use of, health care services, and therefore, may not represent the general population of Hawaiʻi. Individuals that are unable or unwilling to see a health provider may be more likely to have undiagnosed hypertension, especially those without other conditions that may prompt health services utilization.

While the percentage of patients flagged in this sample for undiagnosed hypertension was relatively low (2.7%), the high prevalence of hypertension in the general population translates to a high absolute number of cases without an appropriate diagnosis. Applying an estimate of 2.7% to a population of approximately 1.1 million adults in Hawaiʻi,^23^ 30,000 people would have undiagnosed hypertension and could benefit from hypertension management. As discussed above, this is an underestimate of the true prevalence of undiagnosed hypertension in the population and highlights the need to improve detection.

Our findings underscore the value of leveraging EHR algorithms in identifying patients with potentially undiagnosed hypertension and highlight several factors associated with the lack of diagnosis that can be used to improve diagnosis of hypertension, the leading contributor to death in the US. Our quality improvement evaluation provides evidence that health systems may leverage HIT in identifying, preventing, and managing patients with hypertension.

## Data Availability

The data underlying this article are contained in the electronic health record with use restrictions based on the data use agreement between by the Queen's Health System (QHS) and the Hawai'i Department of Health (HDOH), Chronic Disease Prevention & Health Promotion Division. Data are not publicly available. However, a limited data set may be available from the authors upon reasonable request and with permission of QHS.

## ACKNOWLEDGMENTS

The data underlying this article are contained in the electronic health record with use restrictions based on the data use agreement between by the Queen’s Health System (QHS) and the Hawaiʻi Department of Health (HDOH), Chronic Disease Prevention & Health Promotion Division. Data are not publicly available. However, a limited data set may be available from the authors upon reasonable request and with permission of QHS.

## Non-Standard Abbreviations and Acronyms

US: United States
CDC: Centers for Disease Control and Prevention
HIT: Health Information Technology
EHR: Electronic Health Records
NH: Native Hawaiians
OPI: Other Pacific Islanders
QCIPN: Queen’s Clinically Integrated Physician Network
QHS: Queen’s Health System
HDOH: Hawaiʻi Department of Health
CMS: Centers for Medicare and Medicaid Services
BCBS: Blue Cross Blue Shield
RR: Relative Risk
CI: Confidence Intervals
BMI: Body Mass Index
cRR: Crude Relative Risk
aRR: Adjusted Relative Risk

